# Inferring Respiratory Disease Biology from Geolocation Data

**DOI:** 10.64898/2026.03.05.26347578

**Authors:** Alejandra Rincón Hidalgo, Andrzej K. Jarynowski, Marlli Zambrano, Philip El-Duah, Janik Suer, Ashish Thampi, Richard Pastor, Huynh Thi Phuong, Sten Rüdiger, Stephan Ludwig, Rafael Mikolajczyk, Christian Drosten, Veronika K. Jäger, André Karch, Vitaly Belik, Steven Schulz

## Abstract

Biological fitness quantifies the efficiency and selective advantage of pathogens and hosts in their bilateral interaction. Key questions—such as how much more infectious an emerging variant is compared with its predecessor, or how much protection vaccination offers relative to no vaccination—require fitness to be measured systematically, in real time, and ideally beyond controlled laboratory settings. We propose an approach that infers biological fitness from mostly non-biological data on infection dynamics and contact levels in a population. Because contact levels are expected to predict infection levels under stable biological conditions, systematic deviations between these trends can indicate changes in the underlying processes of transmission or immunity. Using infection surveillance data and GPS co-location information as a proxy for social contact patterns, we apply Bayesian modeling to detect and quantify biological shifts throughout the SARS-CoV-2 pandemic in Germany. We identify substantial regional variation in both the timing and magnitude of fitness changes. These shifts align with the emergence of major variants and the accumulation of population immunity; across Germany, the Alpha, Delta, and Omicron variants were 29 %, 63 %, and 108 % more transmissible than wild-type SARS-CoV-2, respectively. Early natural infection, primary vaccination, and booster vaccination increased population-level immunity by 13 %, 94 %, and 114 % relative to the pre-pandemic naïve state. Our approach provides an immediately deployable framework for detecting biological change during emerging epidemics, given that relevant behavioral and surveillance data are collected in real time. It is particularly valuable for low-resource settings where direct biological measurements may be limited.

**Significance Statement:** Biological changes in pathogen transmission remain difficult to measure in real time during acute epidemic and pandemic situations. Here, we showcase a method to extract biological insights from Bayesian modeling of anonymized mobile phone-based GPS-derived contact data and anonymized infection data. We estimate changes in transmissibility associated with virus evolution and vaccination and find agreement with epidemiological, virological and immunological evidence. Increasingly available mobile phone data makes our approach more easily amenable for use in real time. It enables near real-time detection of biological changes in ongoing (e.g. Influenza) and future epidemics and may be especially valuable in low-resource settings.

## I. INTRODUCTION

Airborne infectious agents, such as Influenza virus and SARS-CoV-2, pose significant risks to public health on the global and local scale [1], in particular when data are scarce or difficult to ascertain and a thorough assessment of acute epidemic situations is therefore beyond reach. This is notably the case for a pathogen’s evolutionary trajectory. The course of a pandemic is dynamically shaped by pathogens that undergo evolution on similar timescales as the immunity mechanisms and non-pharmaceutical interventions that try to counter their effect. However, measuring the effects of virus evolution and immunity typically requires experimental molecular biology methodology which is often infeasible for timely responses and operates under the caveat of generalization beyond lab conditions. Assessing viral variants and immune repertoires requires cumbersome sample collection, genotyping, expression in experimental settings and binding/neutralization assays. This approach lacks information about changing real-world selective pressures, other real-world factors not reflected under experimental conditions, and typically offers limited and retrospective temporal resolution to trace viral and immune evolution in a practically useful manner.

Fitness is a relative measure of the selective advantage of certain pathogen variants or host immune responses over others. Upon emergence of mutations which bene-ficially alter e.g. the binding interface between pathogen and receptor for increased chances of infection or between antibody and epitope for increased chances of neutralization, fitter variants sweep through a host population while purging less fit variants. For pathogens, this so-called fixation process is typically accompanied by transient phases during which several variants are simultaneously present in the host population, also depending on their relative fitness. In the case of SARS-CoV-2, this has been the case notably for the Alpha, Delta and Omicron variants.

The ability to regularly and consistently monitor the evolutionary trajectory of a pathogen-host interaction in terms of fitness with high precision is of enormous interest in virology, immunology, epidemiology and public health. Additionally, timely results are essential for epidemic forecasting and informed interventions [2]. For instance, the emergence of a new variant with increased fitness can rapidly increase the number of new infections and could require new public health interventions. Upticks or drops in fitness of an actively spreading virus could indicate and directly quantify the emergence of fitter variants or the effectiveness of vaccines, respectively, or other changing circumstances which require immediate attention from virologists, immunologists and epidemiologists. Fitness as a representation of a pandemic’s evolutionary trajectory could serve as a valuable indicator of pandemic trends if it could be quantified with high temporal precision.

To quantify fitness in the absence of biological methodology, we leverage technically easily collected contact and infection data as well as the mathematical theory of spreading dynamics in host populations that relate contact and infection numbers to the transmission probability per contact, i.e. transmissibility, which is in turn determined by fitness. This connects “macroscopic” observables (contact and infection numbers) and “microscopic” biological characteristics of disease transmission via mathematical theory and data which is readily available through large-scale, continuous data collection in a population. To estimate transmissibility and fitness, we employ a Bayesian modeling and inference framework and independently collected contact and infection numbers. As a key application case for this methodology, we use the COVID-19 pandemic as the most systematically mapped pandemic to this day: We measure epidemiological, virological and immunological characteristics of SARS-CoV-2 transmission in Germany and their evolutionary dynamics over time. Our methodology provides valuable biological insight both *a posteriori*, as we showcase here, and *in situ* in potential future acute epidemic or pandemic situations if applied to current, systematically recorded contact and infection data. Beyond merely complementing established data and methodology rooted in molecular biology, our approach can provide a more direct path to assess evolving pathogens and immune responses and thus guide public health policy towards informed decision-making in future pandemics.

## II. METHODS

### Base Model

The daily reproduction number *R*_*t*_ quantifies the average number of new infections per infected individual. From a network theoretical perspective, *R*_*t*_ can be related to the mean excess number of contacts *C*_*t*_ and the transmissibility *T*_*t*_ as [3]

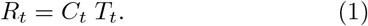

All quantities are defined for specific times denoted by the index *t*. While *C*_*t*_ represents the contribution of contacts to infection dynamics, *T*_*t*_ encompasses the bi-ological aspects of transmission, such as infectiousness – the fitness of the pathogen – and immunity – the fitness of the host (see Fig. 1A). Intuitively, the probability of infection per contact *T*_*t*_ multiplied with the (effective) number of contacts *C*_*t*_ yields the number of new infections *R*_*t*_ (Fig. 1B). While *C*_*t*_ can vary due to changes in contact behavior, *T*_*t*_ varies through changes in viral transmission efficiency. Importantly, it has been noted that epidemic growth does not scale linearly with mean contact numbers ⟨*k*⟩, where *k* denotes individual contact numbers and ⟨·⟩ denotes population average. Instead, because of a network property named *friendship paradox*, they scale with the so-called mean excess number of contacts *C*_*t*_ = ⟨*k*^2^ ⟩*/* ⟨*k*⟩ . It reflects the disproportionate contribution of individuals with a large number of contacts (superspreaders) to the disease dynamics [3–6].

**FIG. 1.**
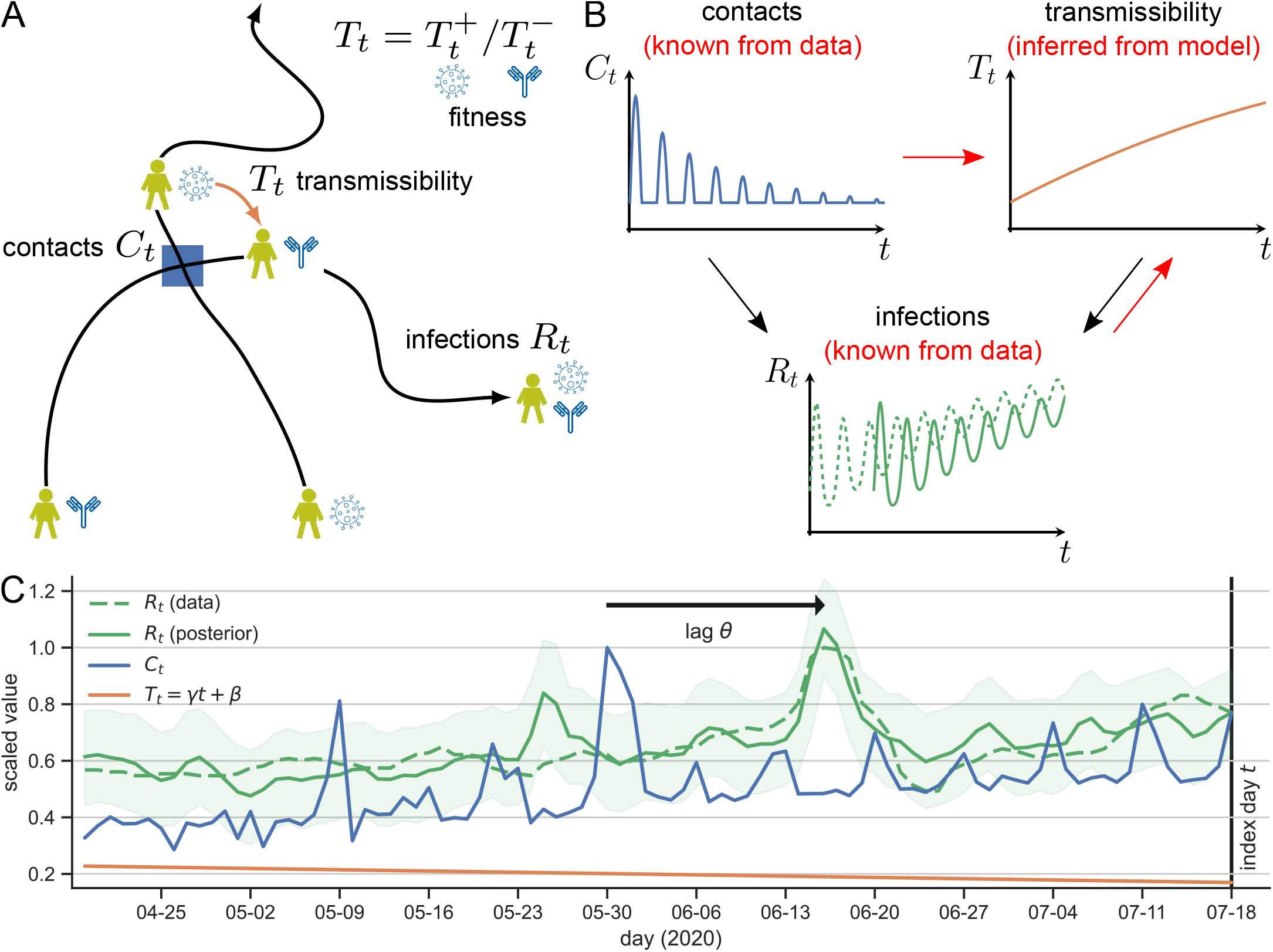
Decomposing infections into contact-driven and biology-driven components using Bayesian modeling. **(A)** Schematic representation of the forward process: Close-range contacts *C*_*t*_ between infectious and susceptible individuals (blue square) multiplied with the transmissibility *T*_*t*_ (orange arrow), yields the number of new infections per infected individual, i.e. the reproduction number *R*_*t*_, *R*_*t*_ = *C*_*t*_*T*_*t*_ (Eq. (1)). Transmissibility *T*_*t*_ itself is decomposed into contributions from viral 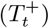 and immune 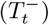 fitness. **(B)** Contacts *C*_*t*_ (blue line) and transmissibility *T*_*t*_ (orange line) can vary over time, here depicted as timeseries, due to changes in social activity (*C*_*t*_) or virus evolution and immunity (*T*_*t*_). Virus evolution is a slow process compared with daily changes in contact activity (the orange line evolves more slowly than the blue line). Black arrows indicate the forward process in (A) which yields the timeseries of infections *R*_*t*_ (green line), typically including a delay from incubation, diagnosis and reporting times (dashed vs. solid green line). Red arrows indicate that transmissibility *T*_*t*_ can be inferred when contacts *C*_*t*_ and infections *R*_*t*_ are known from data. **(C)** Decomposition of the effective reproduction number *R*_*t*_ in the 80-day time window running up to the index day July 18, 2020 in Germany as an example. Scaled original data for daily reproduction numbers *R*_*t*_ (dashed green line) and contacts *C*_*t*_ (blue line) are shown alongside the fitted transmissibility *T*_*t*_ = *γt* + *β* (orange line) and lag *θ* between contact and positive case reporting (black arrow). Posterior *R*_*t*_ (solid green line) with its 90 % credible interval (shaded green) aligns with original *R*_*t*_ data.

The key insight of Eq. (1) is that the “hidden” processes of virus evolution and immunity entering through *T*_*t*_ are connected to the “visible” processes of infection (*R*_*t*_) and contacts (*C*_*t*_) in a simple way. This makes transmissibility *T*_*t*_ accessible from independent measurements of both *R*_*t*_ and *C*_*t*_. In this approach, *T*_*t*_ incorporates all contributions that determine the number of infections besides contact behaviour and therefore integrates the biological fitness of the host-pathogen interaction, including virus evolution, host immunity, and other, e.g. seasonal effects.

### Scaling

We use a log-transformation to turn the multiplicative relationship between the reproduction number *R*_*t*_, contacts *C*_*t*_ and transmissibility *T*_*t*_ in Eq. (1) into an additive relationship. Further, we rescale all quantities to be of *𝒪* (1), i.e. *R*_*t*_ *→ ⟨R⟩*(1 + *R*_*t*_) around the mean value *⟨R⟩*, and similarly for *C*_*t*_ and *T*_*t*_ around *⟨C⟩* and *⟨T ⟩*, respectively. Applying both transformations yields

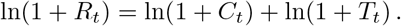

Assuming small values of *R*_*t*_, *C*_*t*_ and *T*_*t*_, i.e. small variations around the rescaling variables ⟨*R*⟩, ⟨*C*⟩ and ⟨*T*⟩, we can use a Taylor expansion to the first order to simplify ln(1 + *x*) *≃ x*. Thus, we obtain

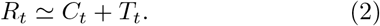

For simplicity, *R*_*t*_, *C*_*t*_ and *T*_*t*_ refer to the rescaled quantities throughout this manuscript.

### Lag structure and linearization

To disentangle the contact and fitness components of infection dynamics, two further adaptions of Eq. (2) are required: First, infections (*R*_*t*_) are typically recorded at the time of a positive test result, which trails the time of the infectious contact (*C*_*t*_) by several days, determined by the incubation period, testing and reporting delays. We account for this temporal lag between the recording of infections and the contact number via a response-delay kernel, i.e. *C*_*t*_ *→ K*_*α,θ*_(*C*_*t*_).

Second, biological processes reflected in *T*_*t*_ evolve more slowly than contacts reflected in *C*_*t*_, as evidenced by empirical studies [7, 8]. Daily variations in contacts are often driven by weekends, holidays, or other short-term factors such as mass events. Computing *T*_*t*_ independently on a day-by-day basis would let daily contact fluctuations confound slow biological trends. We incorporate this separation of timescales by restricting Eq. (2) to short time windows of *W* days, i.e. *t* ∈ [*τ − W* + 1, *τ*[. Here, *τ* denotes the “current” day up to which data for *R*_*t*_ and *C*_*t*_ are available. The slow evolution of biological processes allows to linearize *T*_*t*_ using a constant change rate *γ* within these short time windows, i.e. *T*_*t*_ *→ γt* + *β*.

Applying both changes, we obtain

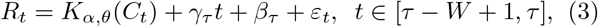

which completes our Bayesian hierarchical model (Fig. S1). Here, *ε*_*t*_ denotes random noise and indices *τ* indicate that the model parameters are inferred independently for every new “current” day *τ* as time proceeds and the newest data for *R*_*t*_ and *C*_*t*_ becomes available. As a delay kernel *K*_*α,θ*_(*C*_*t*_) =∑ _*𝓁≥*0_ *w*_*α,θ,𝓁*_*C*_*t−𝓁*_, we use the Gaussian lag–response operator with weights 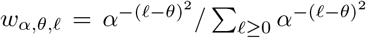. This yields an average lag between contact and now-cast infection of *θ* days with standard deviation of (2 ln *α*)^*−*1*/*2^ days. Eq. (3) describes the instantaneous relationship at time *τ* between *R*_*t*_, *C*_*t*_ and the efficiency change rate *γ*_*τ*_ of biological processes involved in infection. Contacts *C*_*t*_ act as a predictor for the observed target variable *R*_*t*_ and the linear trend term acts as a collector for slow “unobserved” components of infections not explained by *C*_*t*_.

We confirm that the linearization step used to obtain Eq. (3) insignificantly affects the conclusions drawn from a case study in which we apply our methodology to SARS-CoV-2 in Germany (see below): While extending Eq. (3) by the quadratic term in the expansion of *T*_*t*_ improves model performance, it hardly contributes any additional biological insight compared with the linear case (Fig. S3B).

### Transmissibility component

Further insights can be obtained from the model when looking at the transmissibility component in more detail. We can use daily slopes *γ*_*τ*_ of the transmissibility component in Eq. (3) to obtain a (smoothened) measure of transmissibility through trapezoid integration,

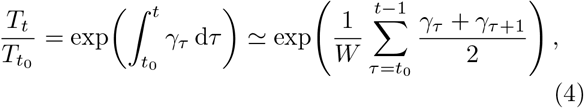

where the exponential reverses the logarithm applied to obtain Eq. (2) from Eq. (1). Due to the prior scaling of the data, the transmissibility measure is defined relative to its value at some baseline time *t*_0_.

Further, we can decompose this transmissibility measure into the individual fitness effects arising from intrinsic host-to-host transmission efficiency of the considered pathogen 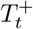 (regardless of population immunity) and from intrinsic immunity of the host population 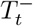 (regardless of the pathogen’s immune escape). These separate periods of monotonously increasing (*γ*_*τ*_ *>* 0) and decreasing (*γ*_*τ*_ *<* 0) transmissibility as immune escape and immunity acquisition, respectively. Mathematically, we can achieve this separation by adapting Eq. (4) to only include the contribution of the periods with positive and negative slopes *γ*_*τ*_,

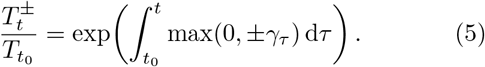

Note that this yields positive values for both 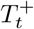 and 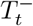. The separation resides on a number of further simplifying assumptions: First, we assume that the composites of *T*_*t*_ can be factored and that immunity acquisition 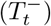 and immune escape 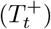 occur sequentially rather than concurrently. Second, 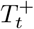 and 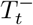 represent effec-tive population-averaged fitness incorporating all sources of transmissibility increase or reduction. Lastly, we assume that pathogen evolution occurs strictly towards increased fitness, i.e. monotonously increasing 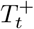 and 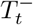, even though neutral and even fitness-decreasing mutations have non-zero takeover probability in finite and spatially structured populations [9, 10].

### Infection dynamics data

We apply our methodology described above to the practical use case of the COVID-19 pandemic in Germany. We utilize official infection surveillance data published by Robert Koch Institute (RKI) for Germany at-large [11, 12] and at the federal state level [13], which provides daily estimates of new infection numbers per infected individual in Germany. Estimates of 7-day now-cast reproduction numbers *R*_*t*_ are computed as the ratio of 7-day average new infections 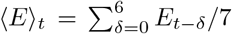 and 7-day average cur-rent infections *⟨E⟩*_*t−*4_ (assumption of a generation time of 4 days [12]), i.e. *R*_*t*_ = ⟨*E*⟩_*t*_*/* ⟨*E*⟩_*t−*4_. Infection numbers *E*_*t*_ are themselves estimates of actual infection numbers accounting for infections on day *t* reported on later days due to incubation, diagnosis and reporting delays (now-casting). While national *R*_*t*_ data for Germany is directly provided by RKI, we computed regional *R*_*t*_ data from infection numbers as described here.

### Contact data

To measure daily mean excess number of contacts *C*_*t*_, we use a GPS-based mobile phone dataset. These data are collected independently from official infection surveillance and cover a sizable sample of the German population. GPS-based mobile phone data allow to identify potentially infectious contacts between phone users as instances of co-location as per their GPS positions at the same time [14, 15]. While such datasets are relatively new, they provide snapshots of a population from a bird’s-eye perspective and have been repeatedly used to study movement and contacts as well as to reveal patterns relevant to spreading dynamics in a population [16–18]. For instance, mobile phone data has been used to assess human mobility patterns and their impact on the spread of SARS-CoV-2 [19]. Moreover, mobile phone data was also used to evaluate non-pharmaceutical interventions (NPIs) such as movement restrictions and physical distancing measures [20]. Our dataset consists of regular individual GPS locations from roughly 1 % of the population in Germany over the course of the entire SARS-CoV-2 pandemic. From these data, we identify contacts and compute individual daily contact numbers *k* as described previously [15] and compute daily mean excess numbers *C*_*t*_ = ⟨*k*^2^⟩*/* ⟨*k*⟩ as a predictor of infections. Here, averages ⟨·⟩ are taken over 1 % of the population and scaled to account for invisible contacts involving the remaining 99 % of the population [15].

### Bayesian inference

Parameter inference is carried out in a Bayesian framework following [21] (Fig. S1), including a set of priors for model parameters to guide inference towards realistic value ranges (Tab. S1). In our retrospective study of SARS-CoV-2 in Germany here, we iterate the “current” day *τ* over the course of the pandemic and fit a new model of the form of Eq. (3) for each choice of *τ* . For every new “current” day *τ*, all parameters of Eq. (3) are estimated from data for *R*_*t*_ and *C*_*t*_ within the last *W* = 80 days running up to day *τ,t* ∈ [*τ* −*W* + 1, *τ*[. This collection of models captures instantaneous relationships between infections, contacts and transmissibility at any given time during the pandemic. For each index day and model, daily infection levels (posterior *R*_*t*_) are sampled from the inferred Bayesian model via the No-U-Turn Sampler (NUTS) with 4 chains, each run with 1000 warm-up and 2000 post-warm-up draws. These are then compared with original data (solid green vs. dashed green lines in Fig. 1C). Convergence diagnostics satisfied 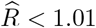 for all parameters, and effective sample sizes exceeded 1000. Trace plots were visually inspected to confirm stationarity and mixing. Identical prior specifications, sampler settings, and convergence diagnostics were used for all days *τ* and all studied regions.

The choice of *W* = 80-day time windows balances trend detection (more accurate at lower *W*) and stable posteriors (more precise at higher *W*) and can be interpreted as a typical timescale of biological trend changes in a pandemic context. Inferred values for the model parameter *γ*_*τ*_, here used to study transmissibility (*T*_*t*_) and fitness (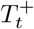 and 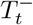), are insensitive to alternative choices for *W* (Fig. S3A).

## III. RESULTS

### Model interpretation

When applying our methodology to the German COVID-19 pandemic data for the exemplary choice of July 18, 2020 as the “current” day *τ*, i.e. the day up to which data is available, Fig. 1C shows the data (*C*_*t*_ and *R*_*t*_) in the 80-day interval leading up to *τ* alongside the trend parameter *γ*_*τ*_ inferred from fitting the model in Eq. (3) to the data.

This visualization illustrates the non-trivial relationship between overall infection dynamics and intrinsic infectiousness of a pathogen revealed by our modeling. While now-cast SARS-CoV-2 infections increase up to a peak on June 17, 2020 (dashed green line), the intrinsic infectiousness of SARS-CoV-2 is effectively unchanged or even slightly declining over the same 80-day time window (orange line with negative slope *γ*_*τ*_). These contrary trends are explained by contacts (blue line). Since contacts and infections increase on par with each other over the course of 80 days, the net effect from intrinsic infectiousness of the pathogen vanishes or slightly decreases. This implies that increased contacts explain increased infections rather than a new SARS-CoV-2 variant with increased fitness in this particular example.

Iterating over the time span from 2020 until 2022 and repeating the procedure for every new “current” day, we find that 97.5 % of *R*_*t*_ data overall falls within the 95 % credible interval of posterior *R*_*t*_; the Pearson correlation between data and posterior *R*_*t*_ is approx. 0.8. These results underscore the fit quality of the model while disentangling the intricate, hidden relationship between intrinsic infectiousness and overall infections.

### Lag time between contact and infection record

The parameter *θ* in Eq. (3) accounts for the temporal lag between the time of contact and infection (*C*_*t*_) and the time of the official record (*R*_*t*_). This lag is expected from a combination of incubation, now-casting, and reporting and diagnostics times and could vary in response to changing biology of the virus, but also changes in testing and reporting protocols. Across all phases of the SARS-CoV-2 pandemic in Germany, the lag time amounts to 14–18 days (Fig. 2), indicating that changes in *R*_*t*_ reflect contact patterns two weeks earlier (Fig. 1C). This range aligns with the expectation from the cumulative time of incubation period, generation-interval offset embedded in infection surveillance as well as additional delays between symptoms and official reporting documented for Germany (see Tab. 1). Additionally, we observe a clear pattern for different phases of the SARS-CoV-2 pandemic in Germany. In 2020, the lag of *θ* = 16.9 days (IQR: 16.4–17.5) is in close agreement with earlier German estimates of the exposure-to-report period [15]. The lag drops to about 14.5 and 15.1 days during the Alpha and Omicron waves, respectively, possibly due to shortened incubation time from a fitter virus or, in the case of Alpha, due to shortened reporting times from the availability of over-the-counter antigen testing in Germany. By contrast, the lag is stabilized at around 16.6 days during the Delta wave, possibly because of a strained diagnosis and reporting system in Germany during this phase.

**TABLE 1.** Time lag between contact and infection record. Time lag *θ* (in days) between contact and now-cast infection record from official infection surveillance in various phases of the SARS-CoV-2 pandemic are extracted from the model. Values correspond to the median as well as the inter-quartile range (IQR) over all index days within the indicated phase. The timeline of each pandemic phase is provided in Tabs. S2 and S3. Reference values for the delay from incubation time are extracted from the literature [32–34]. Now-casting of *R*_*t*_ is estimated under the assumption of a generation time of 4 days [12]. Diagnostic and reporting delays are then imputed as the difference between overall lag *θ* and incubation times and now-casting delays.

| phase | lag $\theta$ [days] | | lag [days] from literature | | |
| --- | --- | --- | --- | --- | --- |
|  | median | [Q1, Q3] | incubation | now-casting | reporting/diagnostic |
| natural immunity | 16.9 | [16.4, 17.5] | 6 | 4 | 6.9 |
| Alpha | 14.5 | [13.7, 16.3] | 5 | 4 | 5.5 |
| regular vaccination I | 16.4 | [15.4, 17.5] |  |  |  |
| Delta I | 16.6 | [15.7, 16.9] | 4.5 | 4 | 8.1 |
| regular vaccination II | 16.6 | [15.8, 17.8] |  |  |  |
| Delta II | 16.7 | [16.1, 17.5] | 4.5 | 4 | 8.2 |
| booster vaccination | 16.2 | [15.2, 17.1] |  |  |  |
| Omicron | 15.1 | [14.2, 16.4] | 3 | 4 | 8.1 |

**FIG. 2.**
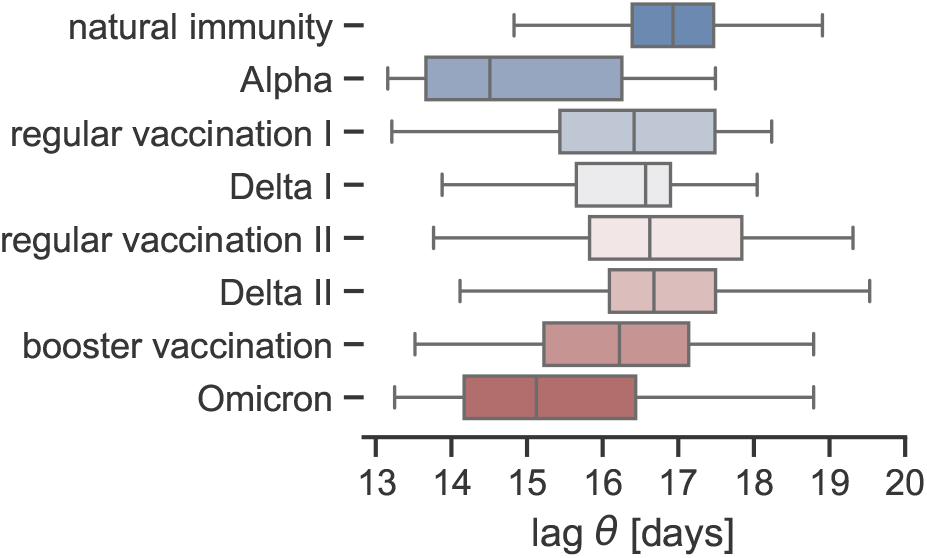
Time lag between contact and infection record. Inferred lag parameter *θ* (in days), measuring the time from infection to infection record in different phases of the SARS-CoV-2 epidemic in Germany. Boxes indicate the posterior median and inter-quartile range (IQR) of *θ* over all index days within each phase, and whiskers extend to farthest values within 1.5 times the IQR. The pandemic phases are defined in Figs. 4 and 5 and start and end dates of pandemic phases are specified in Tabs. S2 and S3. Source data is provided in Tab. 1.

### Virus-host co-evolution

The model parameter *γ*_*τ*_ quantifies diverging trends between infections and contacts, i.e. trends attributable to latent processes other than social contact activity. Constructing the trajectory of *γ*_*τ*_ by iterating the “current” day *τ* over the timeline of the pandemic and fitting the model independently for each *τ* yields the timeseries shown in Fig. S2: When infections (*R*_*t*_) are higher (lower) than expected purely based on contacts (*C*_*t*_) following Eq. (1) in a given 80-day window, we observe positive (negative) slope *γ*_*τ*_ . Periods with negative trends in the latent processes (*γ*_*τ*_ *<* 0) intertwine with periods with positive trend (*γ*_*τ*_ *>* 0).

We use Eq. (4) to compute the (relative) transmissibility *T*_*t*_ of SARS-CoV-2 at every stage of the pandemic in Germany. *T*_*t*_ shows a clear oscillatory behavior across 2021 and 2022, following a constant period near the reference value throughout 2020 (Fig. 3A). This reflects a shift from contact-dominated dynamics in the context of wild-type SARS-CoV-2 and quasi-naïve immunity (2020) to a tug-of-war dynamics between alternating periods of immune escape and immunity acquisition (2021/2022). Strikingly, peaks in *T*_*t*_ align with the emergence of key SARS-CoV-2 variants Alpha, Delta and Omicron (Fig. 3B) while valleys in *T*_*t*_ follow periods of immunization through infection or vaccination (Fig. 3C), thus detecting and quantifying the changing biology of SARS-CoV-2 transmission.

**FIG. 3.**
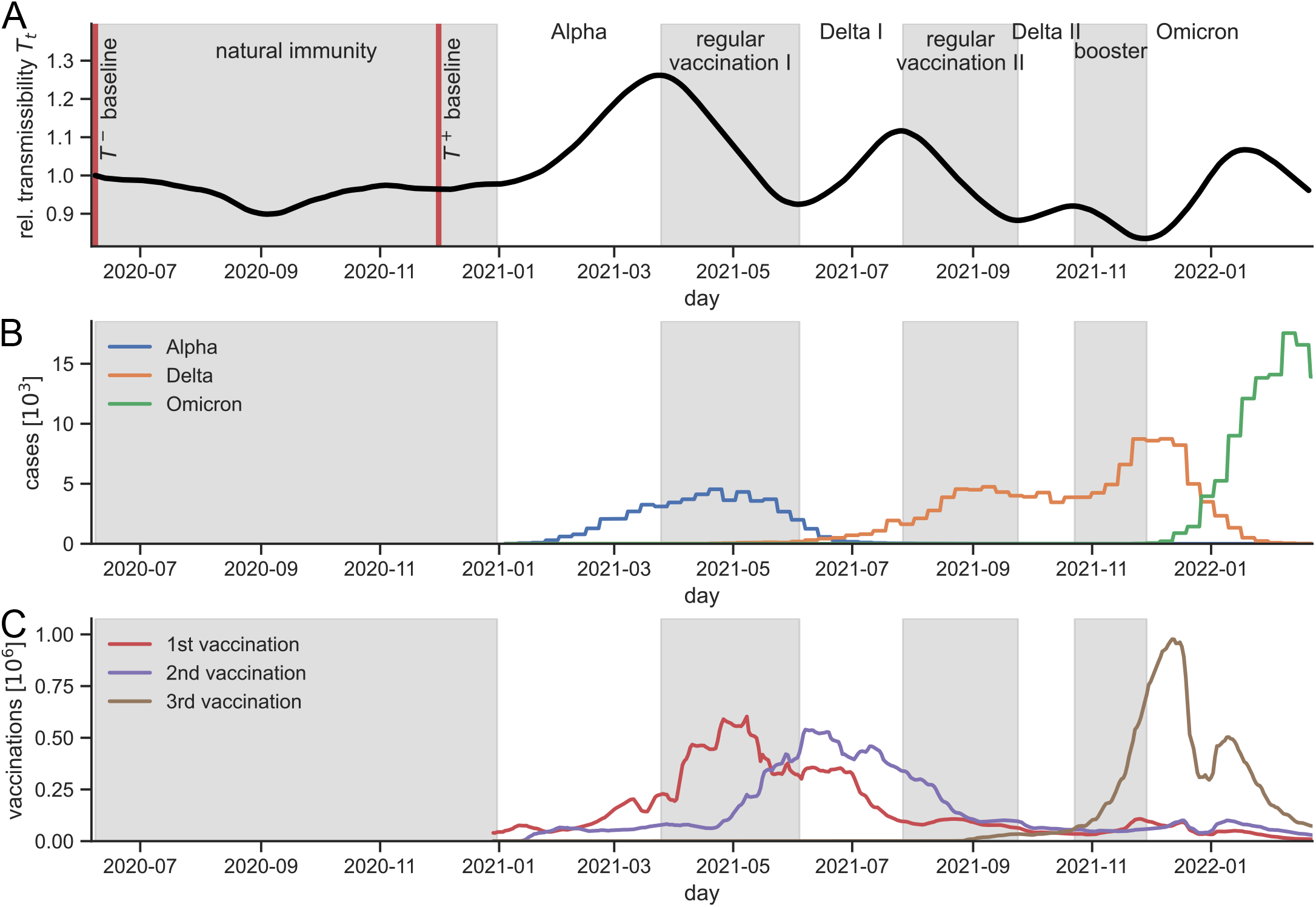
Epidemiological context of SARS-CoV-2 transmissibility *T*_*t*_ in Germany. **(A)** Transmissibility *T*_*t*_ of SARS-CoV-2 in Germany between 2020 and 2022 computed from Eq. (4), relative to the start of the pandemic (*T*_0_ = 1). The epidemiological context of rising and falling trends in *T*_*t*_ is indicated. **(B)** Weekly case numbers by SARS-CoV-2 variant (legend) in Germany. Data taken from [30]. **(C)** 7-day averaged daily vaccination numbers against SARS-CoV-2 in Germany, grouped by the number of vaccines received (legend). Data taken from [31].

To systematically assess whether *T*_*t*_ is the result from the combined use of infection surveillance *R*_*t*_ and contacts *C*_*t*_ and thus reflects an “invisible” latent process not directly captured by the data itself, we build two sets of “negative control” or null models: We infer the model in Eq. (3) as described before, but using “control” data in which contacts *C*_*t*_ within any 80-day interval are either (i) randomly shuffled or (ii) replaced with their mean value (i.e. no contact variation over time). In both cases, we observe that *T*_*t*_ deviates quantitatively and qualitatively from results where contacts are included in the model, especially at times when the effect of contacts was dominant (Fig. S3C). These controls demonstrate that the inference of *T*_*t*_ depends critically on genuine contact data *C*_*t*_ and also that the intrinsic efficiency of biological processes is not reflected by overall infections alone.

### Viral fitness 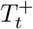

The fitness of SARS-CoV-2 in Germany 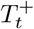 from 2020 through 2022 is shown in Fig. 4A. Phases of increasing 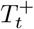 overlap with the takeover of the key SARS-CoV-2 variants Alpha, Delta and Omicron in Germany while intermediate plateaus of constant 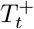 correspond to stable phases where a single variant is dominant in the population. We determine start and end dates of different phases of the SARS-CoV-2 pandemic in Germany from the zero-crossings of *γ*_*τ*_ : Variant takeover phases are initiated when *γ*_*τ*_ changes from negative to positive sign, i.e. 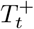 reaches a minimum (Tab. S2). To quantify the fitness increase from key SARS-CoV-2 variants, we compare ratios of 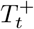 between subsequent plateaus (Tab. S2) and with respect to wild-type SARS-CoV-2 (Fig. 4B, Tab. S2). We find that Alpha, Delta and Omicron had 29 %, 63 % and 108 % increased fitness than wild-type SARS-CoV-2, respectively. These results are in agreement with the range of values in epidemiological (Tab. 2, Fig. 4B) and virological (Tab. 3) studies across various countries.

**TABLE 2.**
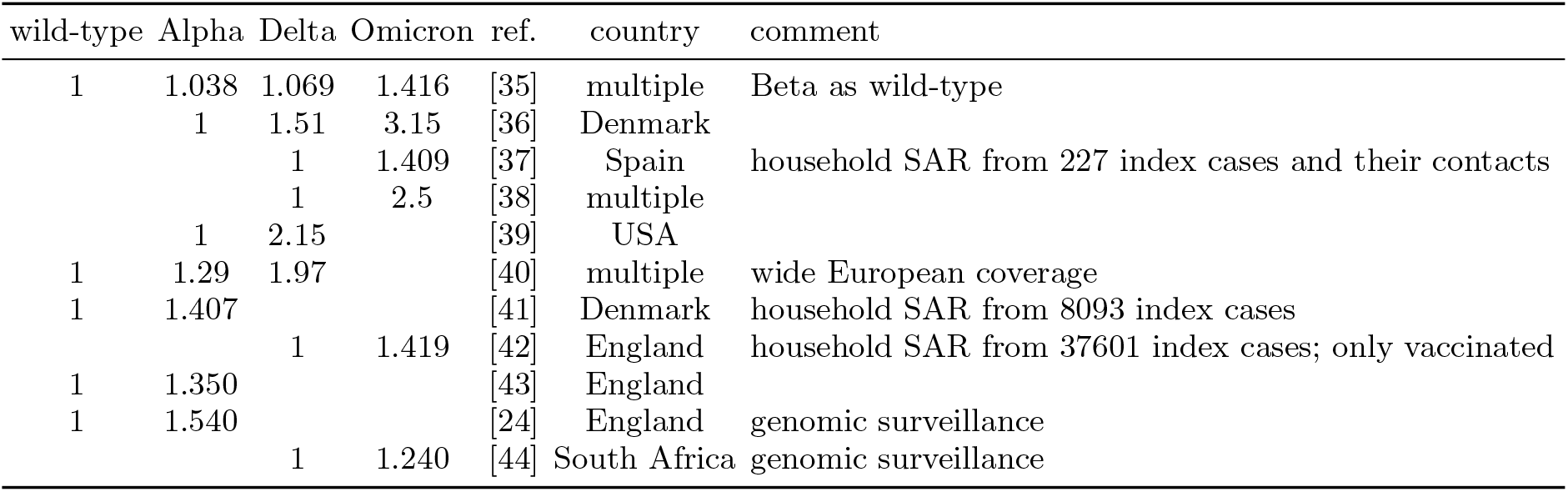
Comparison of key SARS-CoV-2 variants in the epidemiological literature. Each row in the table provides reference to a different epidemiological study or data source. Values indicate by how much each SARS-CoV-2 variant improved in terms of reproduction number compared with the previous variant and are given as ratios with respect to the reference variant (which has a value of 1 in each row). Source data for red dots in Fig. 4B.

**TABLE 3.**
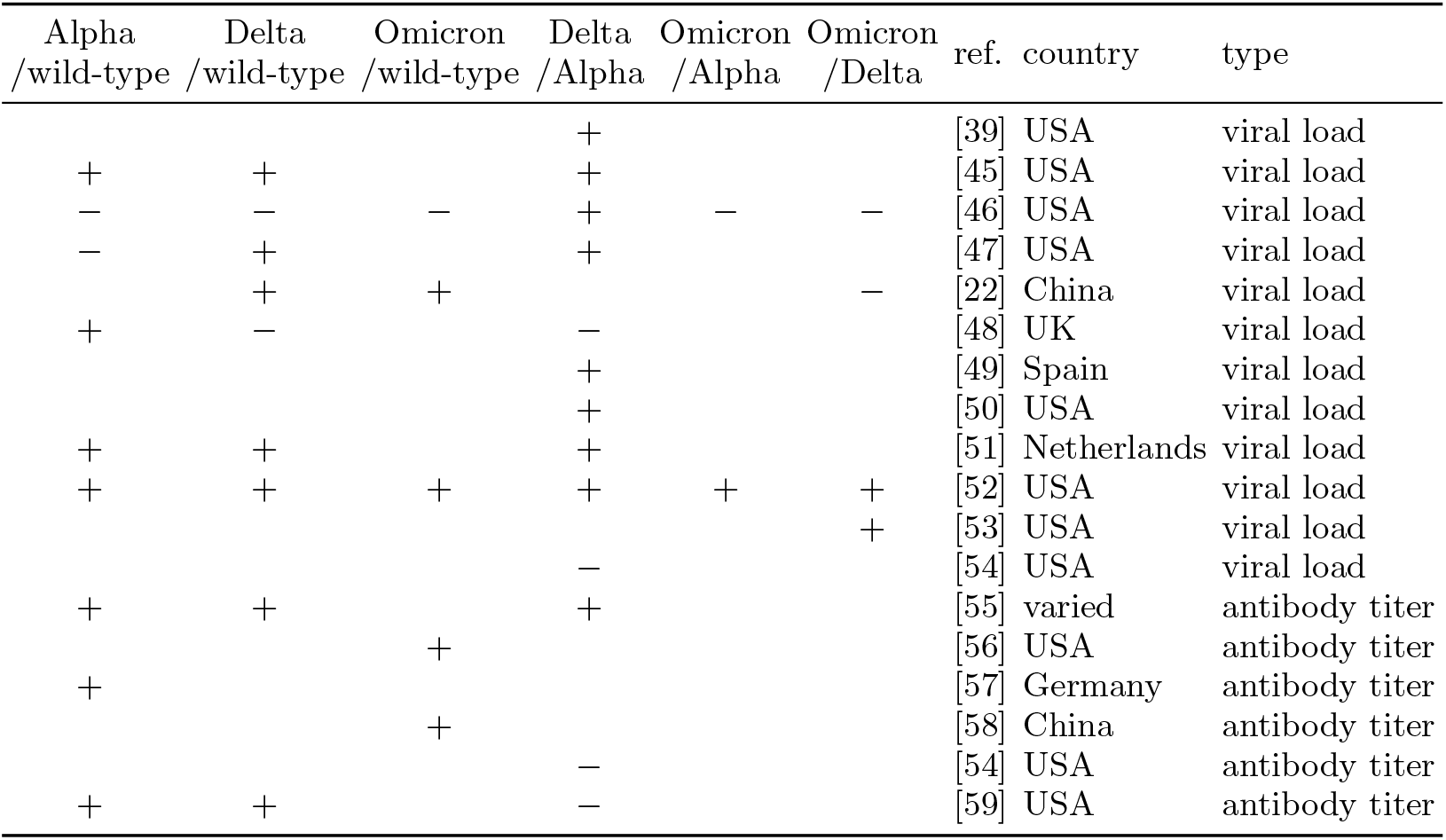
Directional summary of viral load and antibody titer measurements of fitness across SARS-CoV-2 pairs of variants. Each row in the table provides reference to a different virological study or data source; the data type used for comparison (viral load or antibody titer) is indicated. In the columns, each pair of key SARS-CoV-2 variants (wild-type, Alpha, Delta, Omicron) are compared relative to each other, whether the study found increased or decreased viral load/antibody titer: “+” indicates that the subsequent variant is fitter than the preceding one; “*−*” indicates otherwise.

**FIG. 4.**
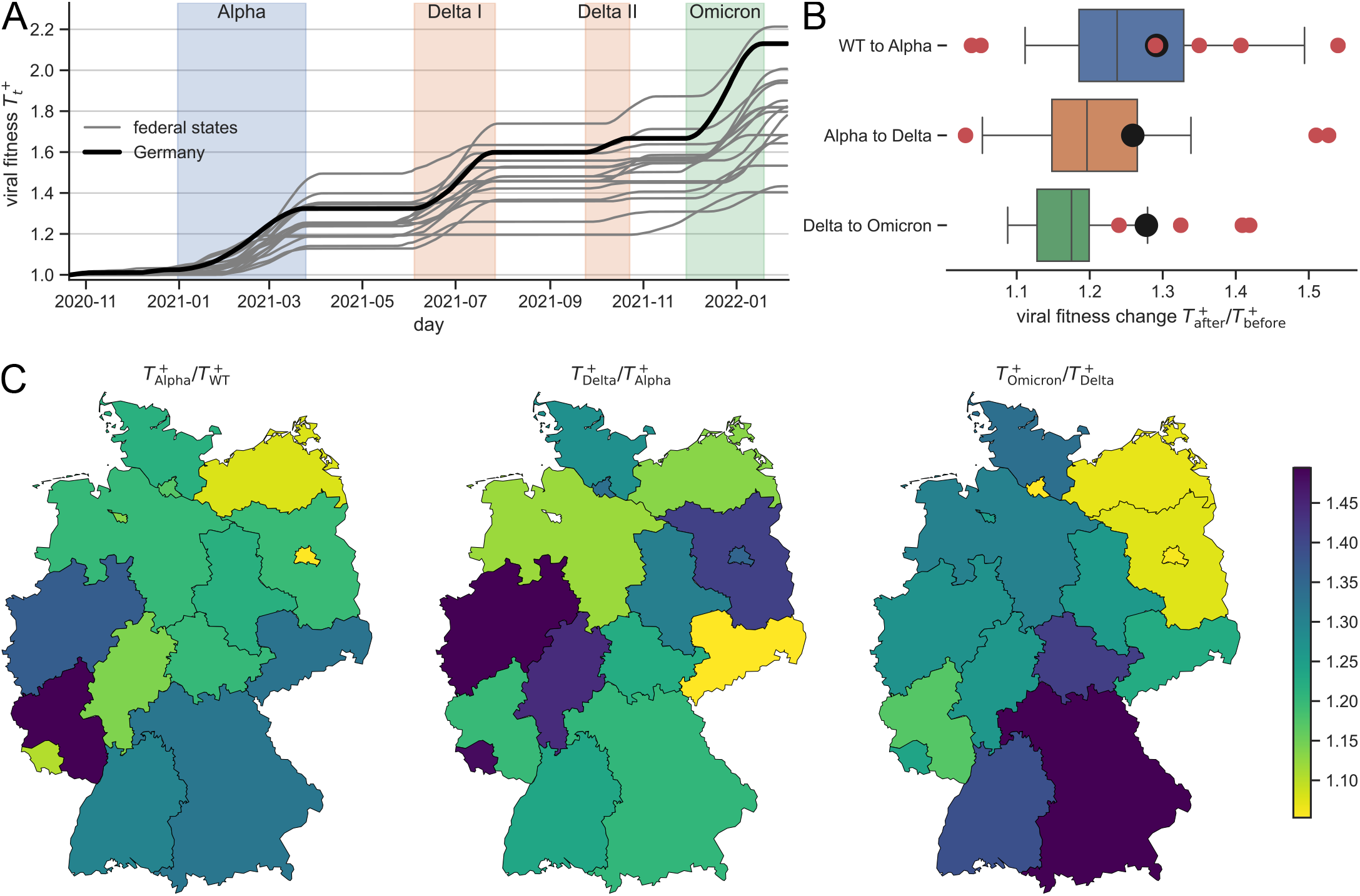
Viral fitness over time and by federal state in Germany. **(A)** SARS-CoV-2 fitness 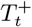 computed from Eq. (5) as a function of time, relative to wild-type SARS-CoV-2 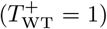, for all 16 federal states in Germany (gray lines; see (C)) and Germany at-large (black line). Step-like rises of 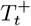 mark the sweeps of key SARS-CoV-2 variants (as labeled) and the timing is highlighted for Germany at-large (shaded time windows). **(B)** Fitness gains 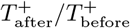 of key SARS-CoV-2 variants compared with the previous key variant expressed as fold-change. Boxes represent the distribution over the 16 German federal states, while black dots represent Germany at-large. Red dots represent reference data from epidemiological studies (Tab. 2). **(C)** For each fitness step in (A), fitness gains 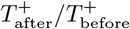 between subsequent SARS-CoV-2 variants are shown at the federal state level in Germany. Source data is provided in Tab. S2.

### Immune fitness 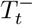

While 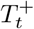 measures viral fitness, 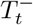 quantifies the fitness of the counteracting force, i.e. immunity against the virus. Measuring the immunological status of a population is an intricate task and subject to many complexities: Immunologically, fitness corresponds to “neutralization” by specific antibodies – the capacity to fend off the virus – which is determined from blood titers from typically small and localized cohorts and is titer- and serotype-dependent. By contrast, we interpret our measure 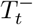 as a simple, population-averaged measure of intrinsic immunity against generic attacks from the same virus, regardless of the antigenic fingerprint and the specificity of immune responses.

We observe five phases of population immunity gain against SARS-CoV-2 in Germany between 2020 and 2022 (Fig. 5A): During 2020, immunity is uniquely acquired through infection with wild-type SARS-CoV-2 (natural immunity) in the absence of vaccines and before the emergence of immune escape variants. Later phases of 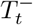 increases are explained by the combined effect of natural immunization with SARS-CoV-2 immune escape variants and active immunization through vaccination campaigns. Notably, the large-scale roll-out of the initial and booster vaccination campaigns are followed by periods of sustained 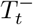 increase (Fig. 3C). To quantify the timing and immunity effects of each wave, we proceed similarly to 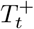. We find that natural immunity against SARS-CoV-2 acquired during 2020 led to 13 % improvement against the quasi-naïve, pre-pandemic status across Germany, compared with 94 % and 114 % improvement against the quasi-naïve status following the initial and booster vaccination campaigns, respectively (Tab. S3, Fig. 5B).

**FIG. 5.**
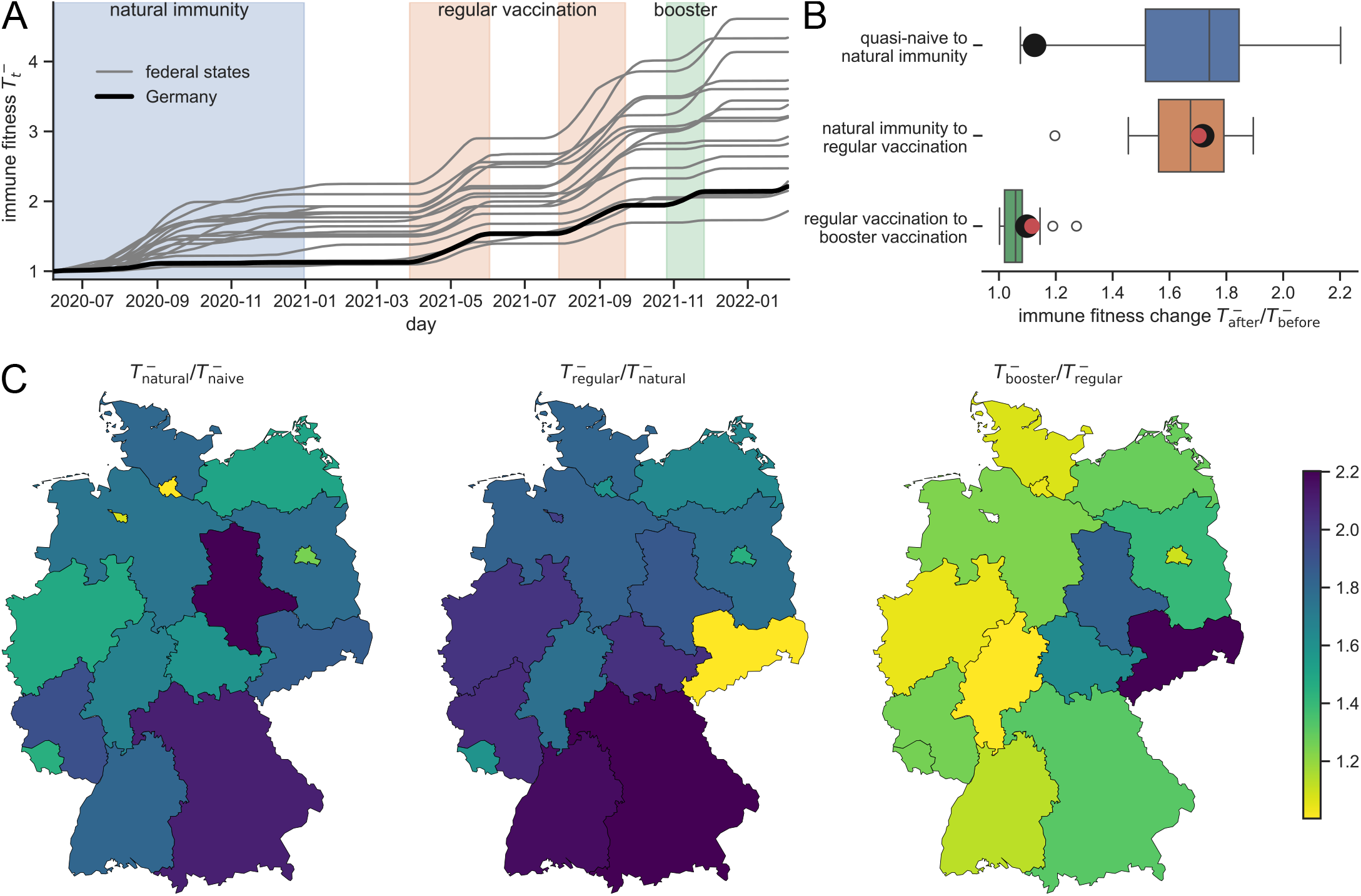
Immune fitness over time and by federal state in Germany. **(A)** Host fitness 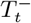 from immunity against SARS-CoV-2 computed from Eq. (5) as a function of time, relative to the quasi-naïve state 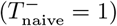], for all 16 federal states in Germany (gray lines; see (C)) and Germany at-large (black line). Step-like rises of 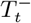 mark phases of immunity acquisition (as labeled) and the timing is highlighted for Germany at-large (shaded time windows). **(B)** Fitness gains 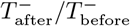 of immunization phases compared with the previous phase expressed as fold-change. Boxes represent the distribution over the 16 German federal states; black dots represent Germany at-large. Red dots represent reference values from antibody levels in blood titers [22]. **(C)** For each fitness step in (A), fitness gains 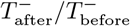 between subsequent immunization phases are shown at the federal state level in Germany. Source data is provided in Tab. S3.

### Comparison with literature

For SARS-CoV-2, we compare our results against data from the epidemiological 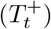, virological 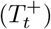 and immunological (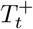 and 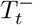) literature and find substantial agreement (Figs. 4B and 5B), thus validating the usefulness of our method.

We extract changes in basic reproduction number (*R*_0,after_*/R*_0,before_) between subsequent SARS-CoV-2 key variants from various epidemiological studies (Tab. 2) to compare our observed viral fitness changes 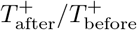. Despite covering various countries besides Germany, the range of literature values coincides with the range of ob-served 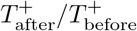 across Germany (Fig. 4B). A com-parison for 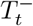 is harder to achieve, as it requires comparing actual antibody levels between patients of different immunity classes on the population level. Immunological studies, however, are typically limited to small, local cohorts. Additionally, such data appears to have been reported only for a single cohort in Shenzhen, China [22], but the SARS-CoV-2 vaccines primarily used in China and Germany bear different mechanisms (whole-virus vs. spike protein epitopes [23]), thereby inducing different immune response patterns. Still, we find surprising agreement when comparing our observed immune fitness changes 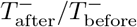 between immunity classes with the corresponding changes in 28-day average IgG antibody levels after symptom onset from [22] (Fig. 5B).

### Spatial heterogeneity

Regional patterns for 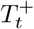 and 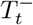 are expected beyond average trends [24], both in timing and in magnitude of the fitness increase phases:Immune fitness may vary e.g. due to unequal vaccine uptake [25] and infection history in the local population [26], whereas viral fitness may depend e.g. on locally varying clonal interference between co-existing viral strains [10] and wave-like expansion from the time and location of first introduction in Germany. Remarkably, the 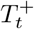 and 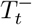 trajectories are almost in lock-step across all 16 German federal states (Figs. 4A and 5A), thus showing that virus evolution and immunity against SARS-CoV-2 had nationwide, nearly simultaneous effects. However, we observe non-trivial regional differences at the level of the German federal states, both in amplitude and timing of the pandemic phases. It is important to note that such fine-grained regional analysis may easily be confounded by local biases in the data (see Discussion).

For instance, 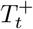 first rises in the adjacent states of Niedersachsen and Schleswig-Holstein as early as November 2020 due to the appearance of the Alpha variant B.1.1.7 (Tab. S2, Fig. 4A). In fact, retrospective genomic analyses showed that B.1.1.7 was first recorded in the city of Hannover, Niedersachsen [27]. While the place of first observation is not necessarily the place of the actual index case, our results suggest that the early circulation and spreading of Alpha in Germany could have occurred within this region. While northern Germany records the earliest 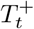 rises, the strongest fitness rise 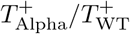 due to B.1.1.7 is observed in the southern states (Tab. S2, Fig. 4C), which is in agreement with the first large-scale B.1.1.7 clusters from genomic analysis appearing in February 2021 in Bavaria [28].

In June/July 2021, first local clusters of the dominant Delta variant B.1.167.2 from genomic analysis in Germany are concentrated in the urban areas of Nordrhein-Westfalen (Cologne, Rhein-Ruhr), Hessen (Frankfurt) and Berlin-Brandenburg, later also in Hamburg [28], which corroborates the corresponding 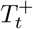 rises in magnitude and in timing (Tab. S2, Fig. 4C). Since the city of Munich is part of the large, mostly rural state of Bavaria, its effect on 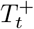 is likely to be shielded in our analysis (Tab. S2, Fig. 4C). Interestingly, B.1.167.2 subsequently sweeps through West Germany while initially sparing East Germany in August/September 2021 before reversing the situation by December 2021/January 2022 [28], following historical boundaries closely. Moreover, vaccine scepticism is more pronounced in East Germany [25]. The combined effect of low vaccine uptake and low infections explains lower 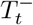 increases in East Germany up to late 2021 (Tab. S3, Fig. 5C), while the region centered around the state of Saxony then undergoes a massive immunity catch-up in December 2021/January 2022 (Tab. S3, Fig. 5C), acquired from both high levels of infection and booster vaccine uptake at this time.

## IV. DISCUSSION

In this work, we describe a new method to characterize “hidden” processes unfolding in a population using GPS co-location data as a proxy for social contacts. Conceptually, while changes in viral and immune fitness occur at the molecular level and are therefore “invisible”, knowledge of the “visible” state of a population (i.e. contacts and infections) enables indirect observation of the “invisible” biology. We showcase the validity of the method by using crowd-sourced GPS co-location contacts as a “bird’s-eye” baseline for SARS-CoV-2 infections. Any trends in actual infections unexplained by contacts are traced to the changing biology of SARS-CoV-2 transmission.

While our method is oriented towards use in potential future epidemic crises, we used the COVID-19 pandemic in Germany as a proof of concept. Our results provide retrospective insights on how viral evolution and immunity jointly sculpt the trajectory of the COVID-19 pandemic in Germany (magnitude, timing and location of fitness hikes due to viral evolution and immunity acquisition). Also, as new variants emerged and population immunity increased at various stages during the COVID-19 pandemic in Germany, the evolving relationship between contacts and infections remained unclear [29]. A major challenge for long-term predictive epidemiological models is reconciling the shifting fitness of pathogens with complex host immune responses. For instance, post-infection and post-vaccination immunity can reduce transmission, whereas waning immunity counteracts these gains by driving infections back up. Meanwhile, viral evolution may introduce more transmissible variants over time, while older variants fade out, thus reducing their impact on overall transmission.

Our estimates of relative changes in variant fitness are quantitatively consistent with epidemiological evidence (Tab. 2, Fig. 4B) and qualitatively consistent with virological studies (Tab. 3). In analytical and field epidemiology, outcomes are typically reported as *R*_*t*_ or as measures that can be mathematically transformed into *R*_*t*_ (e.g., the secondary attack rate, SAR), which enables direct comparison between our estimates and empirical observations. In contrast, virological studies report measurements in units that are not readily convertible into transmissibility. For example, viral-load studies report Ct (cycle threshold) values at detection, log_10_ RNA copies/mL or genome equivalents, and time to viral clearance (the interval between positive test results). Antibody-titer studies use immunoassays e.g., fluorescence/MFI (median fluorescence intensity), BAU (binding antibody units)/mL, AU (arbitrary units)/mL and (ID_50_ neutralization titer giving 50% inhibition) to quantify binding and neutralizing antibodies. Consequently, virological evidence mainly supports the direction of fitness evolution rather than providing directly comparable quantitative estimates, unless international standards are used.

Studying the biology of respiratory disease transmission is of high relevance for public health decision-making. However, typical studies in these fields entail the high organizational and operational cost of cohort recruitment, sample collection and wet-lab processing, e.g. for the study of virus variants, viral load and antibody titers. As a consequence, virological and immunological insights are typically sparse (few time points, little geographical coverage) and are retrospective in nature. By contrast, crowd-based data used in this study can be in principle more easily ascertained and, as we show here, can reveal comparable biological, virological and immunological insights with increased spatio-temporal resolution.

Our work represents a blueprint for the value of anonymous, crowd-based data sources in epidemiological studies. We show that the biology of a contagion can be tracked by means of anonymous population-scale contact and infection data. Compared with typical cohort studies, our method leverages anonymous data donor panels covering a substantial share of the population at the expense of representativeness to allow for statistically significant contact levels within and across populations as a baseline. Besides data size, crowd-based data collection is also automatable and scalable and in principle available in real time, thus enabling a continuous and geographically resolved assessment of transmission biology as we demonstrate in this paper.

Our work sheds new light on the challenge of epidemic forecast: predicting future infections *R*_*t*_ from data of the present. Elaborate methods which use heterogeneous, readily available data sources (mobility data, social network data, climate data, etc.) oftentimes show limited success. The reason may reside in the lack of critical virological and immunological transmission information in near real time and future infections may therefore be intrinsically hard to predict. We here reverse this task by using present infections *R*_*t*_ as input to infer the missing near real-time insights about the biology of disease transmission which could prove more valuable than attempts to accurately predict the future. It is in this respect that our work reconciles independent biology- and non-biology-focused streams of research during the SARS-CoV-2 pandemic.

Our model outputs could prove immediately actionable when applied to real-time data and could be of high relevance for policymakers: Infection trends beyond what is expected from contact trends, reflected in *γ*_*τ*_ and *T*_*t*_, can be monitored regularly akin to weather forecasts and be used to assess the possibility of fitter virus variants and the effectiousness of immunization. When *γ*_*τ*_ deviates significantly from zero, behavioral interventions such as lockdowns alone are unlikely to curb growth, and biological counter-measures (e.g. variant-tailored boosters) should be prioritized. Conversely, a large negative *γ*_*τ*_ signals that existing immunity is suppressing transmission, allowing for the cautious relaxation of contact restrictions.

A substantial limitation of our approach resides in the need for consistent data collection over long periods of time and across different regions. Inconsistencies owing e.g. to protocol changes in infection surveillance or crowd-based data donation represent major sources of bias and can lead to unwanted trends unrelated to the transmission process and false alarms with our method. In particular, the validity of our model depends on reliable daily *R*_*t*_ estimates. Issues with the underlying data could have led to an overestimation of 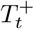 for Alpha and underestimation for Delta in the state of Rhineland Pala-tine (Fig. 4). Also, the extent of our study is limited up to early 2022 (takeover of the Omicron variant in Germany) as infection surveillance protocols changed massively from this time on. For other airborne pathogens such as seasonal Influenza virus and RSV, routine surveillance in Germany is very uneven. Alternative, possibly more systematic incidence proxies such as wastewater surveillance could be less prone to inconsistencies and replace infection surveillance used in this work.

Moreover, geographical resolution is limited by the depth of GPS-based data collection, especially because our method requires the estimation of higher moments of underlying distributions. For the small federal state of Bremen, this could be hardly achieved (see missing signal from Delta variant and booster vaccine in Tabs. S2 and S3). All these shortcomings could be remedied for future use of our method e.g. by government-initiated and narrowly purposed data donation campaigns during acute epidemic crises, which could lead to population shares well beyond 1 %.

## Data Availability

Scripts as well as input and intermediate data files to reproduce all figures and results in this manuscript have been deposited in a public GitHub repository: https://github.com/st-sch/nc-mmm-forecast-gui/tree/for-paper.

## ACKNOWLEDGMENT

A.R.H., A.T., R.P., S.R., and S.S. were supported by the German Federal Ministry for Economic Affairs and Climate Action (BMWK) via the project DAKI-FWS (grant number 01MK21009A). A.K.J., M.Z., J.S.,H.T.P., R.M., V.K.J., A.K., and V.B. were supported by the German Federal Ministry for Education and Research (BMBF) via the project OptimAgent (grant number 031L0299A). Additionally, A.K.J., M.Z., and V.B. were partially supported by “Pandemic non-pharmaceutical interventions to flatten the curve: needs, effectiveness and impact in the global South – the example of Ghana”, primarily funded by the Berlin University Alliance (BUA) as part of the Excellence Strategy of the German Federal and State Governments (grant number 113 MC GlobalHealth). M.Z. and V.B. were also partially supported by the German Federal Ministry of Research, Technology and Space (BMFTR) via the project DREAM-EP (“Data-informed Responsive Epidemic Analysis and Multiscale-Modelling for Epidemic Preparedness”) within the MONID – Phase II – Consortium (grant number 031L0323E).

## ETHICS STATEMENT

The Ethics Committee of the Medical Board Westfalen-Lippe and the University of Münster gave ethical approval for this work (reference number 2020–473-f-s).

Participation in this opt-in study was voluntary, and an informed consent was obtained from each of the participants. All analyses were carried out on anonymised data. The co-location contact data are collected via a software development kit developed for the primary purpose of assessing the quality of cell phone networks. The general terms and conditions of this data collection (www.netcheck.de/datenschutz) also cover the use for research purposes through a broad consent. All users have agreed to the data collection by opt-in. Data collection and usage in this work was reviewed and approved by law firm GvW Graf von Westphalen (gvw.com), deemed compliant with regulations under Federal German Law with regard to protection of privacy and personal information (DSGVO).

## AUTHOR CONTRIBUTIONS

Conceptualization: A.R.H., A.K.J., P.E., A.T., R.P.,S.R., S.L., V.B., S.S. Data curation: A.R.H., A.K.J.,P.E., A.T., R.P., S.R., S.S. Formal analysis: A.R.H.,A.K.J., M.Z., V.B., S.S. Funding acquisition: R.P., S.R.,R.M., V.K.J., A.K., V.B. Investigation: A.R.H., A.K.J.,M.Z., P.E., J.S., R.P., H.T.P., S.R., S.L., V.B., S.S.Methodology: A.R.H., A.K.J., R.P., S.R., V.B., S.S.Project administration: A.K.J., R.P., S.R., R.M., C.D.,V.K.J., A.K., V.B., S.S. Software: A.R.H., A.K.J., A.T.,V.B., S.S. Resources: A.R.H., A.K.J., M.Z., P.E., A.T.,R.P., C.D., V.B., S.S. Supervision: A.K.J., R.M., C.D.,V.K.J., A.K., V.B., S.S. Validation: A.R.H., A.K.J.,M.Z., J.S., H.T.P., S.R., S.L., R.M., C.D., V.K.J., A.K., V.B., S.S. Visualization: A.R.H., A.K.J., V.B., S.S. Writing of the draft: A.R.H., A.K.J., M.Z., J.S., S.L., V.K.J., A.K., V.B., S.S.

## COMPETING INTERESTS

A.R.H., A.T., R.P., and S.S. are employees of NET CHECK GmbH. S.R. is a former employee of NET CHECK GmbH. M.Z. is an intern at NET CHECK GmbH. All other authors declare no competing interests.

---

**FIG. S1.**
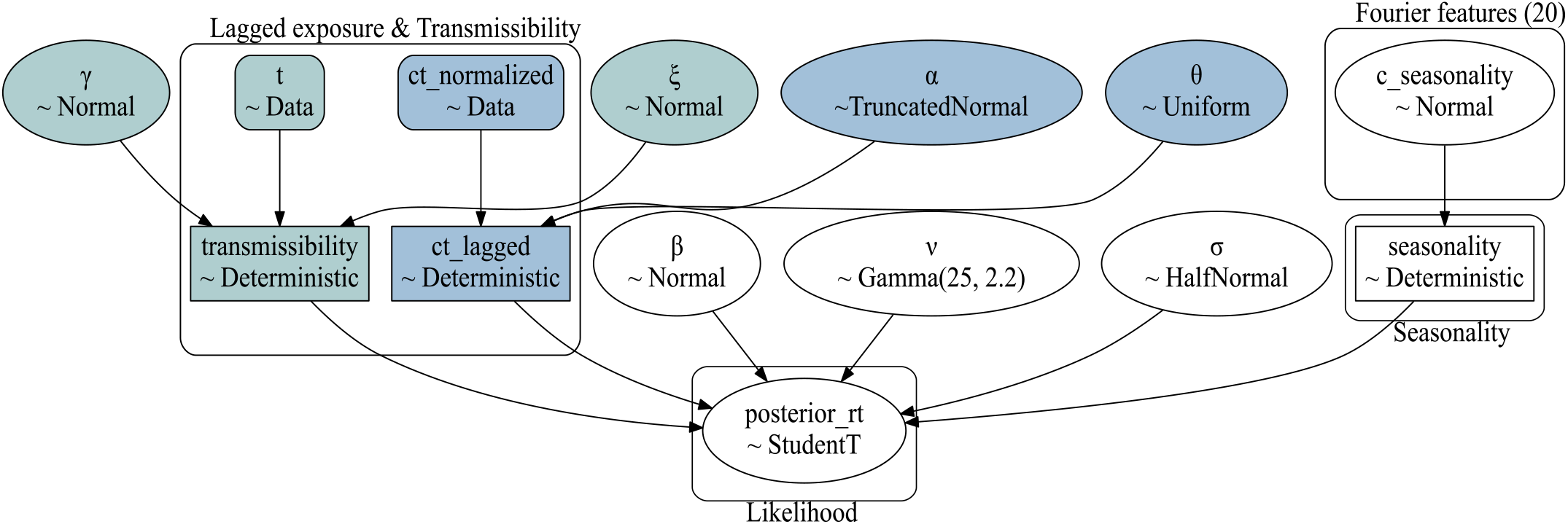
Schematic of the Bayesian model. Model parameters and prior distributions for the Bayesian decomposition of *R*_*t*_ (Eq. (3)) are illustrated. Parameters highlighted in blue govern the Gaussian-lagged contact response *K*_*α,θ*_(*C*_*t*_), while parameters highlighted in green control contributions from linearized transmissibility *T*_*t*_ = *γt* + *β*. Rectangles denote deterministic transformations or observed data streams.

**FIG. S2.**
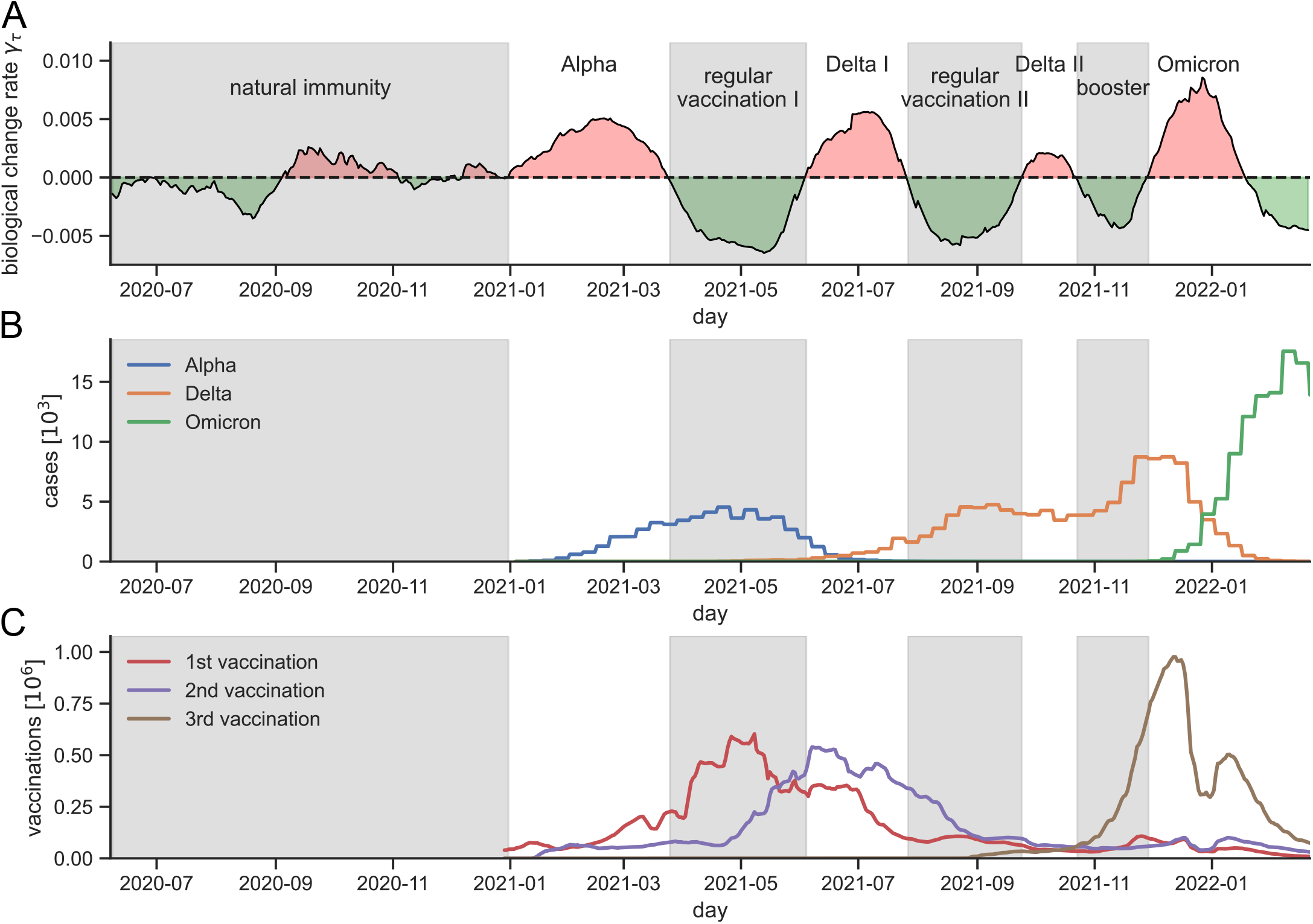
Relation between the trend parameter *γ*_*τ*_ of SARS-CoV-2 and the emergence of VOCs and vaccines in Germany. **(A)** Evolution of the trend parameter *γ*_*τ*_ for SARS-CoV-2 over time between 2020 and 2022 in Germany as determined from the Bayesian modeling. For each index day *τ*, the Bayesian model is re-estimated on data for the *W* = 80 days running up to *τ* and the posterior mean of the slope parameter *γ*_*τ*_ is recorded. Pandemic phases identified from the zero-crossings of *γ*_*τ*_ are highlighted. **(B)** Weekly number of infections identified with specific SARS-CoV-2 variants of concern (VOC) in Germany [30]. **(C)** 7-day-averaged daily vaccination numbers against SARS-CoV-2 in Germany, grouped by the number of vaccines received (legend) [31].

**FIG. S3.**
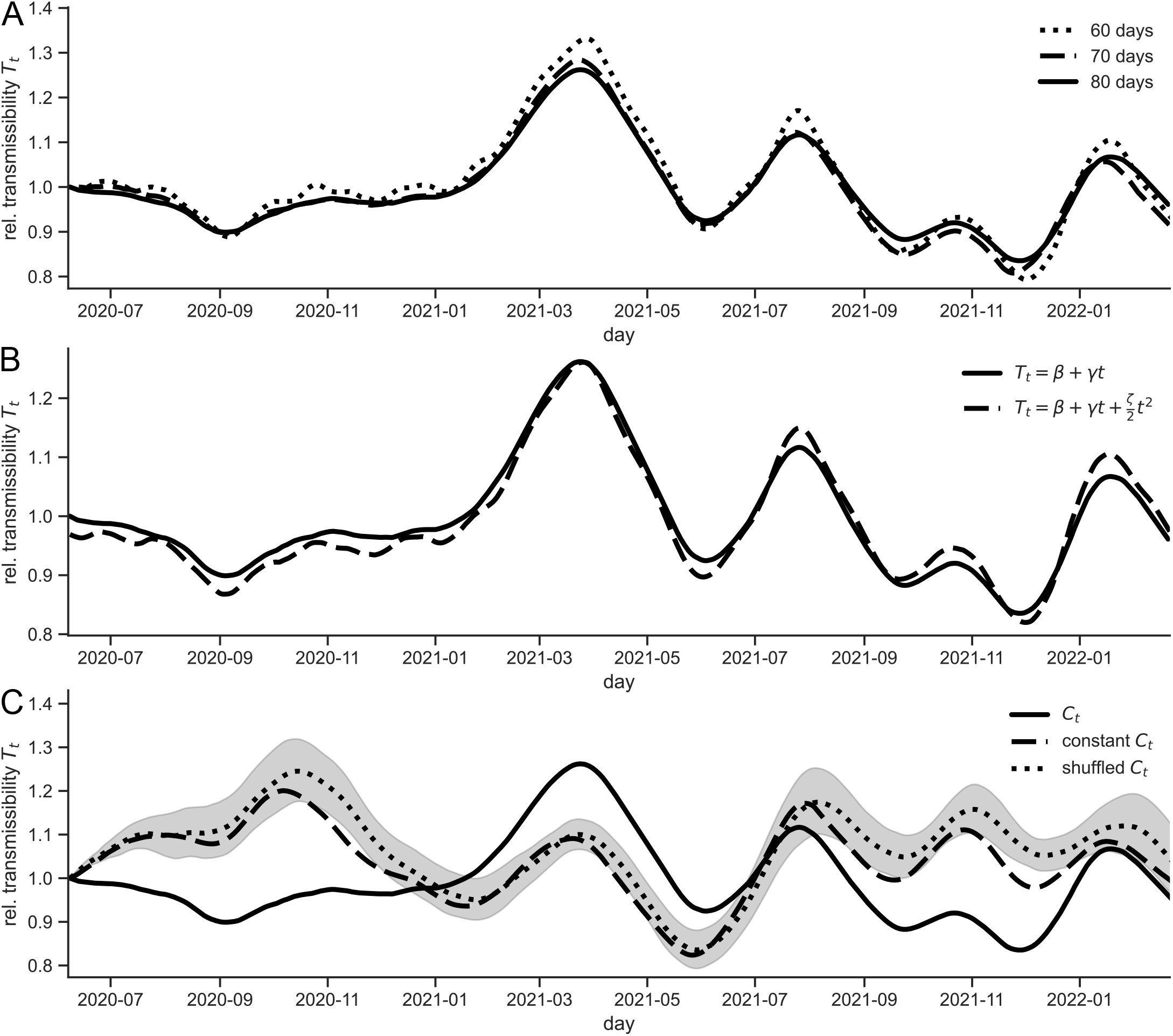
Various control cases for the inference of SARS-CoV-2 transmissibility *T*_*t*_ in Germany. **(A)** Comparison of *T*_*t*_ computed from Eq. (4) for multiple choices of the data time window *W* used for model training. **(B)** Comparison of *T*_*t*_ between the linear approximation of *T*_*t*_ within time windows of *W* days and inclusion of the quadratic term. In the quadratic case, the integral equation to compute *T*_*t*_ (Eq. (4)) is extended according to the Euler-Maclaurin formula, i.e.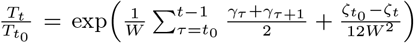. **(C)** Comparison of *T*_*t*_ against null models in which the data for contacts *C*_*t*_ had been replaced by a constant value (i.e. its mean value over the time window of *W* days) or randomly shuffled among the *W* days. The grey-shaded area shows the 95 % confidence interval over 10 independent realizations of the data shuffling. In all panels, the transmissibility signal *T*_*t*_ for further analysis throughout this manuscript (which uses genuine contact data *C*_*t*_ at the Bayesian inference step and the linear approximation for *T*_*t*_ within *W* = 80-day time windows) is shown as solid black line.

**TABLE S1.** Parameter specification of the Bayesian model. Interpretation and prior distributions of parameters in the Bayesian regression model (Eq. (3) and Fig. S1). The truncated normal distribution𝒩 ^[*a,b*]^(*µ, σ*) is confined to the interval [*a, b*]. Seasonality is modeled with 20 Fourier terms, thus sharing the same weakly–informative prior.

| parameter | prior | interpretation and hyper-parameters |
| --- | --- | --- |
| $\gamma_\tau$ | $\mathcal{N}(0, 1)$ | local slope of transmissibility $T_t$ |
| $\theta_\tau$ | $\mathcal{U}(\theta_1, \theta_2)$ | peak lag of Gaussian kernel $K_{\alpha_\tau, \theta_\tau}(\cdot)$ ($\theta_1 = 13$ days, $\theta_2 = 21$ days) |
| $\xi_\tau$ | $\mathcal{N}(0, 1)$ | local intercept of transmissibility $T_t$ |
| $\alpha_\tau$ | $\mathcal{N}^{[0, 0.4]}(0.2, 0.1)$ | retention rate ($0 \leq \alpha \leq 0.4$) |
| $\beta_\tau$ | $\mathcal{N}(1, 0.015)$ | scaling of delayed effect of contacts $K_{\alpha_\tau, \theta_\tau}(C_t)$ |
| $c_{\text{season}, k}$ | $\mathcal{N}(0, 0.025)$ | Fourier coefficients ($k = 1, \dots, 20$) for weekly seasonality |
| $\varepsilon_t$ | $\mathcal{T}(0, \sigma_\tau^2, \nu_\tau)$ | residual noise |
| $\sigma_\tau$ | HalfNormal(0.5) | scale parameter of Student-$t$ likelihood $\mathcal{T}$ |
| $\nu_\tau$ | Gamma(25, 2.2) | degrees of freedom parameter of Student-$t$ likelihood $\mathcal{T}$ |

**TABLE S2.** Fitness 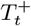 of SARS-CoV-2 and regional variations. For each German federal state and Germany at-large (rows) as well as each key SARS-CoV-2 variant (columns), the start and end dates of the fitness step as well as the fitness gain compared with the previous variant 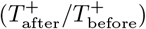 and with wild-type SARS-CoV-2 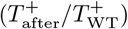 are given as fold-changes. Missing values indicate that no fitness increase could be detected. Source data for Fig. 4.

|  | Alpha |  |  |  | Delta |  |  |  | Omicron |  |  |  |
| --- | --- | --- | --- | --- | --- | --- | --- | --- | --- | --- | --- | --- |
| | start | end | $\frac{T_{\text{Alpha}}^+}{T_{\text{WT}}^+}$ | $\frac{T_{\text{Alpha}}^+}{T_{\text{WT}}^+}$ | start | end | $\frac{T_{\text{Delta}}^+}{T_{\text{Alpha}}^+}$ | $\frac{T_{\text{Delta}}^+}{T_{\text{WT}}^+}$ | start | end | $\frac{T_{\text{Omicron}}^+}{T_{\text{Delta}}^+}$ | $\frac{T_{\text{Omicron}}^+}{T_{\text{WT}}^+}$ |
|  | 2020/21 |  |  |  | 2021 |  |  |  | 2021/22 |  |  |  |
| Baden-Wuerttemberg | 01-12 | 04-03 | 1.323 | 1.323 | 06-13 | 11-05 | 1.170 | 1.548 | 12-03 | 01-31 | 1.233 | 1.908 |
| Bayern | 12-26 | 04-04 | 1.345 | 1.345 | 06-01 | 07-19 | 1.151 | 1.547 | 12-05 | 01-31 | 1.279 | 1.979 |
| Berlin | 02-01 | 03-27 | 1.111 | 1.111 | 06-03 | 10-31 | 1.246 | 1.385 | 11-27 | 01-13 | 1.090 | 1.510 |
| Brandenburg | 01-26 | 03-27 | 1.235 | 1.235 | 05-19 | 07-17 | 1.284 | 1.586 | 12-12 | 01-25 | 1.097 | 1.739 |
| Bremen | 12-15 | 02-26 | 1.183 | 1.183 | – | – | – | – | 10-22 | 01-08 | 1.174 | 1.389 |
| Germany | 12-31 | 03-25 | 1.292 | 1.292 | 06-04 | 10-23 | 1.259 | 1.626 | 11-29 | 01-19 | 1.278 | 2.077 |
| Hamburg | 01-29 | 04-01 | 1.213 | 1.213 | 06-09 | 11-03 | 1.235 | 1.498 | 12-01 | 01-10 | 1.088 | 1.629 |
| Hessen | 12-18 | 03-11 | 1.186 | 1.186 | 05-21 | 11-08 | 1.302 | 1.544 | 11-26 | 01-22 | 1.178 | 1.819 |
| Mecklenburg-Vorpommern | 01-31 | 03-28 | 1.133 | 1.133 | 05-22 | 07-09 | 1.104 | 1.250 | 12-12 | 01-31 | 1.095 | 1.369 |
| Niedersachsen | 11-30 | 03-29 | 1.237 | 1.237 | 05-30 | 07-15 | 1.096 | 1.356 | 12-04 | 01-26 | 1.195 | 1.621 |
| Nordrhein-Westfalen | 12-18 | 04-05 | 1.382 | 1.382 | 06-06 | 11-09 | 1.339 | 1.850 | 11-28 | 01-26 | 1.182 | 2.187 |
| Rheinland-Pfalz | 12-18 | 04-04 | 1.494 | 1.494 | 05-27 | 11-06 | 1.146 | 1.712 | 12-11 | 01-31 | 1.138 | 1.948 |
| Saarland | 01-14 | 03-31 | 1.155 | 1.155 | 05-29 | 11-10 | 1.330 | 1.537 | 12-11 | 02-01 | 1.167 | 1.794 |
| Sachsen | 01-23 | 04-01 | 1.348 | 1.348 | 06-18 | 07-26 | 1.053 | 1.420 | 12-23 | 02-07 | 1.162 | 1.650 |
| Sachsen-Anhalt | 01-28 | 03-29 | 1.253 | 1.253 | 05-23 | 07-16 | 1.216 | 1.524 | 12-23 | 02-08 | 1.176 | 1.792 |
| Schleswig-Holstein | 11-29 | 03-27 | 1.251 | 1.251 | 05-20 | 10-28 | 1.196 | 1.496 | 11-21 | 01-20 | 1.210 | 1.811 |
| Thuringen | 01-13 | 04-01 | 1.238 | 1.238 | 05-26 | 07-20 | 1.162 | 1.439 | 12-20 | 02-15 | 1.243 | 1.789 |

**TABLE S3.** Host fitness 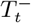 and regional variations from natural immunization and vaccination. For each German federal state and Germany at-large (rows) as well as each immunization step (columns), the start and end dates of the fitness step as well as the fitness gain compared to before 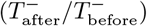 and with the pre-pandemic quasi-naïve state 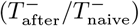are given as fold-changes. Missing values indicate that no fitness increase could be detected. Source data for Fig. 5.

|  | natural immunity |  |  |  | regular vaccination |  |  |  | booster vaccination |  |  |  |
| --- | --- | --- | --- | --- | --- | --- | --- | --- | --- | --- | --- | --- |
| | start | end | $\frac{T_{\text{natural}}^-}{T_{\text{naive}}^-}$ | $\frac{T_{\text{natural}}^-}{T_{\text{naive}}^-}$ | start | end | $\frac{T_{\text{regular}}^-}{T_{\text{natural}}^-}$ | $\frac{T_{\text{regular}}^-}{T_{\text{naive}}^-}$ | start | end | $\frac{T_{\text{booster}}^-}{T_{\text{regular}}^-}$ | $\frac{T_{\text{booster}}^-}{T_{\text{naive}}^-}$ |
|  | 2020 |  |  |  | 2021 |  |  |  | 2021 |  |  |  |
| Baden-Wuerttemberg | 06-08 | 01-12 | 1.841 | 1.841 | 04-03 | 10-12 | 1.888 | 3.476 | 11-05 | 12-02 | 1.036 | 3.603 |
| Bayern | 06-08 | 12-26 | 2.101 | 2.101 | 04-04 | 10-13 | 1.910 | 4.013 | 10-29 | 12-05 | 1.079 | 4.329 |
| Berlin | 06-08 | 02-01 | 1.380 | 1.380 | 03-27 | 10-03 | 1.471 | 2.029 | 10-31 | 11-27 | 1.030 | 2.090 |
| Brandenburg | 06-08 | 01-26 | 1.830 | 1.830 | 03-27 | 10-04 | 1.656 | 3.031 | 11-04 | 12-12 | 1.095 | 3.318 |
| Bremen | 06-08 | 12-15 | 1.150 | 1.150 | 03-01 | 10-19 | 1.788 | 2.056 | – | – | – | – |
| Germany | 06-08 | 12-31 | 1.125 | 1.125 | 03-25 | 09-24 | 1.726 | 1.943 | 10-23 | 11-29 | 1.102 | 2.140 |
| Hamburg | 06-08 | 01-29 | 1.097 | 1.097 | 04-01 | 09-27 | 1.544 | 1.694 | 11-03 | 12-01 | 1.021 | 1.728 |
| Hessen | 06-08 | 12-18 | 1.711 | 1.711 | 03-11 | 10-11 | 1.654 | 2.829 | 11-08 | 11-26 | 1.013 | 2.866 |
| Mecklenburg-Vorpommern | 06-08 | 01-31 | 1.565 | 1.565 | 03-28 | 09-27 | 1.581 | 2.475 | 11-06 | 12-12 | 1.067 | 2.641 |
| Niedersachsen | 06-08 | 11-30 | 1.770 | 1.770 | 03-29 | 10-09 | 1.700 | 3.008 | 10-31 | 12-04 | 1.060 | 3.190 |
| Nordrhein-Westfalen | 06-08 | 12-18 | 1.523 | 1.523 | 04-05 | 10-06 | 1.801 | 2.743 | 11-09 | 11-28 | 1.018 | 2.792 |
| Rheinland-Pfalz | 06-08 | 12-18 | 1.927 | 1.927 | 04-04 | 10-09 | 1.817 | 3.501 | 11-06 | 12-11 | 1.063 | 3.722 |
| Saarland | 06-08 | 01-14 | 1.496 | 1.496 | 03-31 | 10-07 | 1.276 | 1.910 | 11-10 | 12-11 | 1.061 | 2.027 |
| Sachsen | 06-08 | 01-23 | 1.928 | 1.928 | 04-01 | 09-25 | 1.206 | 2.326 | 10-21 | 12-23 | 1.280 | 2.978 |
| Sachsen-Anhalt | 06-08 | 01-28 | 2.249 | 2.249 | 03-29 | 10-04 | 1.716 | 3.858 | 11-01 | 12-23 | 1.195 | 4.610 |
| Schleswig-Holstein | 06-08 | 11-29 | 1.828 | 1.828 | 03-27 | 09-29 | 1.679 | 3.069 | 10-31 | 11-21 | 1.025 | 3.145 |
| Thuringen | 06-08 | 01-13 | 1.642 | 1.642 | 04-02 | 10-28 | 1.820 | 2.990 | 10-30 | 12-20 | 1.150 | 3.439 |

## Notes

### Author Declarations

The Ethics Committee of the Medical Board Westfalen-Lippe and the University of Münster gave ethical approval for this work (reference number 2020-473-f-s). Participation in this opt-in study was voluntary, and an informed consent was obtained from each of the participants. All analyses were carried out on anonymised data. The co-location contact data are collected via a software development kit developed for the primary purpose of assessing the quality of cell phone networks. The general terms and conditions of this data collection (www.netcheck.de/datenschutz) also cover the use for research purposes through a broad consent. All users have agreed to the data collection by opt-in. Data collection and usage in this work was reviewed and approved by law firm GvW Graf von Westphalen (gvw.com), deemed compliant with regulations under Federal German Law with regard to protection of privacy and personal information (DSGVO).

